# SLEEP QUALITY AND CIRCADIAN RHYTHM DISRUPTION IN CRITICALLY ILL PATIENTS IN INTENSIVE CARE UNIT

**DOI:** 10.1101/2024.09.10.24313393

**Authors:** Sampani Christina-Athanasia, Koukoulitsios Georgios, Liveri Athanasia, Dimitrios Papageorgiou

## Abstract

**Aim:** The aim of this study was to evaluate the quality of sleep in critically ill patients in the Intensive Care Unit (ICU).

**Method and Material:** This study investigated the night-time sleep of 135 patients admitted to the general ICU of the General Hospital of Athens “G. Gennimatas” between January 2021 and December 2023. Data were collected using the Richards Campbell Sleep Questionnaire (RCSQ). Sensory stimuli, including noise, light, nursing activities, and invasive procedures, were reduced during the night to improve patients’ sleep. Measures to reduce light included implementing special lighting during nursing tasks and using bedside lamps during care. Noise reduction strategies involved closing doors, minimizing monitor alarms, and discouraging staff from speaking near patient beds. Grouping patient care activities was also promoted to limit sleep disturbances.

**Results:** Patients in the intervention group showed improved sleep quality compared to the control group, with a significant overall RCSQ score (p<0.05). Gender and age did not significantly affect sleep quality. However, hospital stay duration differed between groups, with the control group experiencing shorter stays. A negative correlation was observed between the duration of hospitalization and sleep quality, with longer stays linked to lower RCSQ scores. Additionally, extended mechanical ventilation was associated with poorer sleep quality.

**Conclusion:** Patients in the ICU often exhibit abnormal levels of alertness, poor quality of daytime sleep, disrupted nighttime sleep, and sleep patterns that lack both slow-wave and rapid eye movement (REM) sleep. Gaining a deeper understanding of the role circadian rhythms play in managing critical illness could pave the way for future chronotherapeutic strategies, enhancing clinical outcomes and promoting recovery for patients.

## Introduction

Sleep disturbances are a prevalent issue among patients in Intensive Care Units (ICUs), with sleep deprivation and circadian rhythm disruptions recognized as serious complications in critically ill individuals. Poor sleep quality in the ICU is often associated with longer hospital stays, increased mortality rates, and the onset of delirium. The ICU environment itself, mechanical ventilation, medications, and the severity of a patient’s illness are major contributors to disrupted sleep patterns.^1^

Sleep is crucial for restoring the body’s normal functions, with the circadian rhythm playing a key role in regulating the sleep-wake cycle. In ICU patients, disturbances in sleep quality and circadian rhythm are widespread, and these disruptions can have significant effects on patient recovery and overall health.^2^

Critically ill patients frequently experience poor sleep quality due to a range of environmental and physiological factors. Continuous monitoring by staff, bright lighting, and constant noise in the ICU disturb the natural sleep cycle, contributing to sleep deprivation and further complicating patient recovery. Sleep disturbances are often linked to an increased risk of infections, decreased respiratory function, elevated pain levels, and delirium, a common complication among critically ill patients.^3^

The circadian rhythm, which controls the sleep-wake cycle and regulates hormone secretion and other vital functions, is often disrupted in ICU patients. The absence of natural light, along with artificial lighting used during the night, can disturb the body’s biological clock. This disruption leads to irregular sleep patterns, increased fatigue, and a prolonged recovery time. The effects of circadian rhythm disruption in ICU patients are severe, contributing to diminished immune function, higher levels of inflammation, and a worsened overall prognosis.^4^

Certain medications administered in the ICU, such as sedatives, antipsychotics, and opioids, also play a role in affecting sleep quality. Medications like propofol and benzodiazepines are known to suppress critical stages of sleep, particularly REM and NREM stages, which are essential for restoring bodily functions. Prolonged use of these medications can lead to fragmented sleep, and abrupt discontinuation may result in rebound insomnia. Additionally, pain management is crucial for maintaining sleep quality in ICU patients, as inadequate pain relief can cause frequent awakenings. Anxiety, stress, and the inability to communicate due to illness further exacerbate sleep disturbances.^5,6^

Mechanical ventilation is another major factor that disrupts sleep in ICU patients. Patients on mechanical ventilators often experience frequent interruptions due to equipment, tubes, and the effort required for breathing, which reduces total sleep time and sleep quality. In recent years, research has increasingly focused on understanding the relationship between mechanical ventilation and sleep disturbances, highlighting the need for better strategies to mitigate these effects.^7^

Improving sleep quality in the ICU is essential for facilitating faster patient recovery. Strategies such as reducing noise levels, adjusting lighting to mimic natural circadian rhythms, and utilizing non-pharmacological interventions like light therapy and earplugs have been explored as potential solutions. Additionally, optimizing the timing of medication administration and minimizing the use of sedatives can help restore circadian rhythm and improve sleep quality.^8^

Sleep studies in ICU patients, using polysomnography, show that compared to healthy adults, ICU patients experience fragmented sleep, prolonged sleep latency, and reduced sleep efficiency. These studies indicate that approximately 50% of sleep in ICU patients occurs during the daytime and is characterized by transitions to lighter sleep stages. Sleep disturbances in the ICU are multifactorial, influenced by environmental factors such as noise and light, many of which can be modified to improve sleep quality.^9^

Sleep disturbances are widespread among ICU patients, with prevalence rates ranging from 22% to 61% across various studies. Epidemiological data from Europe and the United States highlight the significant impact of factors like noise, frequent staff interventions, and patient anxiety on sleep quality. In Europe, around 50% of ICU patients report sleep problems, while in the United States, about 70% of adults experience poor sleep quality at least once a month.^10,11,12^

In Greece, data on sleep disturbances in ICU patients are limited, primarily coming from small-scale studies or individual hospital reports. Sleep disorders, including insomnia and sleep apnea, are prevalent in the general population, affecting individuals across various age groups and genders. These sleep difficulties are closely associated with physical and emotional problems, mental health disorders, and chronic health conditions such as cardiovascular disease, obesity, and diabetes.^13,14^

Efforts to improve sleep quality in ICU patients, including non-pharmacological interventions like bright light therapy and earplugs, have yielded mixed results. The lack of natural light and excessive artificial light during the night remain key challenges in promoting better sleep and maintaining circadian rhythms in the ICU setting.^15^

## Methodology

### Participants

The aim of this study was to evaluate the quality of sleep among critically ill patients in the Intensive Care Unit (ICU). The study sample consisted of 135 patients admitted to the ICU of the General Hospital of Athens “G. Gennimatas” from January 2021 to December 2023. It was utilized a convenience sampling approach. The ICU facility consisted of 17 beds and maintained a nurse-to-patient ratio of 1:2.

Eligible participants were patients aged 16 years and above, both with and without the need for mechanical ventilation, and exhibiting hemodynamic stability. Exclusion criteria included patients under 16 years of age, those with hemodynamic instability, sedation, a history of sleep-disordered breathing (such as sleep apnea syndrome), chronic neuromuscular disease, psychiatric illness, previous sleep pathologies, alcohol addiction, illicit drug abuse, and cognitive dysfunction (including dementia).

Data collection adhered to strict anonymity and confidentiality protocols. The process commenced only after obtaining informed and voluntary consent from each patient. To maintain integrity and confidentiality, the data in the questionnaires were coded and anonymized. Each patient was assigned a unique code number with no direct reference to their identity.

### Description of Richards-Campbell Sleep Questionnaire (RCSQ)

Data collection was conducted using the Richards-Campbell Sleep Questionnaire (RCSQ), which was completed by the researcher during the study. The RCSQ is a brief, self-reported questionnaire consisting of 5 items used to assess nighttime sleep quality. Specifically, it evaluates:

1. Sleep depth
2. Time to fall asleep
3. Number of awakenings
4. Return to sleep (percentage of time awake)
5. Overall sleep quality

Each item is rated on a visual analogue scale ranging from 0 mm to 100 mm, with higher scores indicating better sleep quality. The average score of the five items is known as the “total score” and represents the overall perception of sleep. Additionally, a sixth item was included to assess Night-time Noise Level (range: 0 mm for “very quiet” to 100 mm for “very noisy”). Demographic and clinical characteristics of the participants (gender, age, length of stay, days on mechanical ventilation, days on spontaneous breathing, whether they underwent tracheostomy and the type of tracheostomy) were also collected.

### Intervention

The intervention in this study was conducted in the ICU. It involved techniques to reduce sensory stimuli (noise, light, nursing activities, invasive procedures) during the night and recorded the quality of sleep of the patients. Measures to reduce light included implementing a lighting program for nursing procedures or conducting night-time care activities with bedside lighting when possible. Noise control measures included closing doors when not in use, reducing alarms from monitors, and adjusting phone volumes. Staff were discouraged from talking around patient beds, and efforts were made to consolidate patient care and treatment activities to minimize the number of individual disturbances for each patient.

### Ethical issues

Regarding the ethics of this study, it has been carried out in accordance with The Code of Ethics of the World Medical Association (Declaration of Helsinki). The study was approved by the hospital’s review boards (Ref No 3369/8-2-2021). Data collection and analysis were conducted after obtaining informed, written consent from all patients’ relatives during ICU care. The patients’ personal data and the hospital’s name remained anonymous at all stages of the study.^16^

### Statistical Analysis

Data analysis was performed using SPSS version 24. Descriptive statistics for quantitative variables were presented as means and standard deviations (M ± SD), while categorical variables were presented as absolute (n) and relative frequencies (%). Normality tests were conducted using the Kolmogorov-Smirnov test. Factor analysis was performed to determine the construct validity of the RCSQ. Data adequacy for factor analysis was assessed using the Kaiser-Meyer-Olkin (KMO) measure and Bartlett’s test of sphericity. Reliability of the RCSQ was assessed using Cronbach’s alpha coefficient (α). Values of the index greater than 0.7 or 0.8 are generally considered satisfactory. Differences between RCSQ scores and demographic-clinical characteristics were explored using parametric t-tests and non-parametric Mann-Whitney tests. Correlations between two quantitative variables were examined using Pearson’s correlation coefficient (r) for parametric and Spearman’s rank correlation coefficient (ρ) for non-parametric data. Statistical significance was set at p<0.05.

## Results

### Demographic Characteristics

The sample of this study consisted of 135 individuals, of whom 71.1% (n=96) were men and 28.9% (n=39) were women, with an average age of 56.70 (SD=16.35).

The mean duration of hospitalization was 39.07 (SD=62.83) days, the mean duration of mechanical ventilation was 34.79 (SD= 62.23) days, and the mean duration of spontaneous respiration was days 4.40 (SD=3.65).

Among the participants, 56.3% (n=76) had undergone a tracheostomy, while 43.7% (n=59) had not (**Table 1**).

**Table 1:** Demographic and Clinical Characteristics of the Sample.

|  |  |
| --- | --- |
| **Gender** |  |
| Male/Female | 71,1% (96)/ 28.9% (39) |
| **Age(years), mean ± SD** | 56.70±16.35 |
| **Days of Hospitalization, mean ± SD** | 39.07±62.83 |
| **Days on Mechanical Ventilation, mean ± SD** | 34.79±62.23 |
| **Days on Spontaneous Breathing, mean ± SD** | 4.40±3.65 |
| **Tracheostomy N (%)** |  |
| No/ Yes | 43.7% (59)/ 56.3% (76) |
| **Type of Tracheostomy** |  |
| Surgical/ Percutaneous | 19.7% (15)/ 80.3% (61) |

### Characteristics of RCSQ

The average score for depth of sleep was 56.00 (SD=15.17), time to fall asleep was 48.37 (SD=18.66), number of awakenings was 46.22 (SD=16.16), return to sleep was 50.96(SD=17.95), sleep quality was 47.19(SD=20.68), and noise level was 51.04(SD=19.29) (**Table 2**). The mean total RCSQ score without the noise level question was 49.759SD=15.09), a moderate score, while the mean total RCSQ score including the noise level question was 49.96(SD=15.60). Of the patients, 48.9% (n=66) reported good sleep, 40.7% (n=55) reported poor sleep, 8.1% (n=11) reported very poor sleep, and 2.2% (n=3) reported very good sleep.

**Table 2:** Patient Scores on the RCSQ Questionnaire.

| Parameter | Mean | SD | Range |
| --- | --- | --- | --- |
| Depth of Sleep | 56.00 | ±15.17 | 0-100 |
| Time to Fall Asleep | 48.37 | ±18.66 | 0-80 |
| Number of Awakenings | 46.22 | ±16.16 | 0-80 |
| Return to Sleep | 50.96 | ±17.95 | 0-80 |
| Sleep Quality | 47.19 | ±20.68 | 0-100 |
| Noise Level | 51.04 | ±19.29 | 0-80 |
| Total RCSQ (5 parameters) | 49.75 | ±15.09 | 6-78 |
| Total RCSQ (including noise level) | 49.96 | ±15.60 | 5-78 |

### Factor Analysis of RCSQ

The adequacy of the data for factor analysis was tested using the Kaiser-Meyer-Olkin (KMO) measure and Bartlett’s sphericity test. The factor analysis showed that the KMO measure was 0.846 and Bartlett’s sphericity test had a chi-square value of 483.670 with p<0.05, indicating that the data is suitable for factor analysis (**Table 3**).

**Table 3:** KMO and Bartlett’s Test.

| Measure | Value |
| --- | --- |
| Kaiser-Meyer-Olkin Measure of Sampling Adequacy | 0.846 |
| Bartlett's Test of Sphericity, Approx. Chi-Square | 483.670 |
| Degrees of Freedom (df) | 10 |
| Significance (p) | <0.001 |

### Cronbach’s Alpha Reliability Coefficient for RCSQ

The reliability of the RCSQ was tested using Cronbach’s alpha coefficient. The coefficient was calculated as 0.900, indicating excellent reliability of the RCSQ. No removal of questions significantly increased the value of the coefficient.

### Comparisons and Correlations between Demographic Characteristics and Patient Group

The gender of the participants did not seem to relate to the patient group (p=0.466). Both the intervention and control groups had more men than women.

The age of the patients did not differ between the control and intervention groups (p=0.881). However, the days of hospitalization appeared to differ between the two groups (p=0.006). Individuals in the control group had fewer days of hospitalization compared to those in the intervention group. Additionally, the days on mechanical ventilation differed by patient group (p=0.07). Patients in the control group had fewer days on mechanical ventilation compared to patients in the intervention group. The days on spontaneous breathing did not differ between the control and intervention groups (p=0.115). No statistically significant relationships were found between the patient group and whether they had undergone a tracheostomy (p=0.703), their transfer (p=0.212), or the receipt of mild sedation for sleep promotion (p=0.391) (**Table 4**).

**Table 4:** Distribution of demographic characteristics between control group and intervention group.

|  | Control Group (n=62) | Intervention Group (n=73) | P value |
| --- | --- | --- | --- |
| **Gender** |  |  |  |
| Male/Female | 74.2% (46)/ 25.8% (16) | 68.5% (50)/ 31.5% (23) | 0.466 |
| **Age** | 57.61 ± 15.21 | 55.65 ± 17.28 | 0.881 |
| **Days of hospitalization** | 37.31 ± 51.38 | 41.04 ± 71.85 | 0.006 |
| **Days on mechanical ventilation** | 32.34 ± 49.67 | 37.29 ± 71.88 | 0.007 |
| **Days on spontaneous breathing** | 4.97 ± 3.99 | 3.90 ± 3.29 | 0.115 |
| **Tracheostomy** |  |  |  |
| No/ Yes | 41.9% (26)/ 58.1% (36) | 45.2% (33)/ 54.8% (40) | 0.703 |

### Comparisons and correlations of demographic characteristics and RCSQ

The participants’ gender did not show differences in the RCSQ scales nor in the overall RCSQ score (p>0.05) (**Table 5**).

**Table 5:**
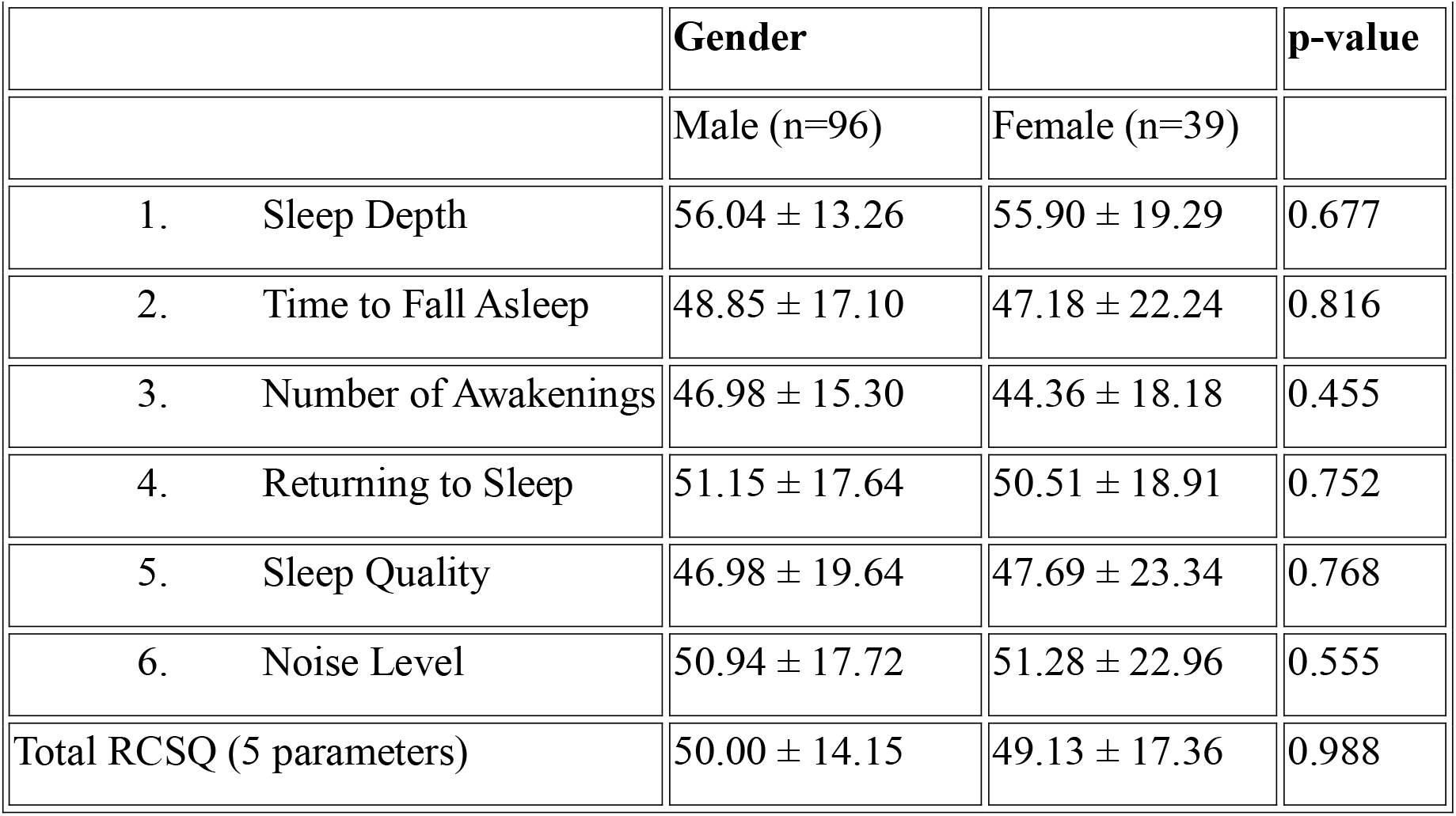
Comparisons between RCSQ and gender.

|  | Gender |  | p-value |
| --- | --- | --- | --- |
|  | Male (n=96) | Female (n=39) |  |
| 1. Sleep Depth | 56.04 ± 13.26 | 55.90 ± 19.29 | 0.677 |
| 2. Time to Fall Asleep | 48.85 ± 17.10 | 47.18 ± 22.24 | 0.816 |
| 3. Number of Awakenings | 46.98 ± 15.30 | 44.36 ± 18.18 | 0.455 |
| 4. Returning to Sleep | 51.15 ± 17.64 | 50.51 ± 18.91 | 0.752 |
| 5. Sleep Quality | 46.98 ± 19.64 | 47.69 ± 23.34 | 0.768 |
| 6. Noise Level | 50.94 ± 17.72 | 51.28 ± 22.96 | 0.555 |
| Total RCSQ (5 parameters) | 50.00 ± 14.15 | 49.13 ± 17.36 | 0.988 |

The age of the patients did not appear to be related to any of the RCSQ scales nor to the total RCSQ score (p>0.05) (**Table 6**).

**Table 6:** Correlations between RCSQ and age.

|  | Age | p-value |
| --- | --- | --- |
| 1. Sleep Depth | 0.067 | 0.439 |
| 2. Time to Fall Asleep | -0.058 | 0.501 |
| 3. Number of Awakenings | -0.013 | 0.878 |
| 4. Returning to Sleep | -0.045 | 0.603 |
| 5. Sleep Quality | -0.073 | 0.403 |
| 6. Noise Level | -0.091 | 0.293 |
| Total RCSQ (5 parameters) | -0.031 | 0.722 |

In contrast, the length of patients’ hospitalization appeared to be negatively correlated with the RCSQ scales and the total RCSQ score (p<0.05). Slight and very slight statistically significant negative correlations were found, suggesting that as the duration of patients’ hospitalization increases, their scores on the RCSQ scales and the total RCSQ score decrease (**Table 7**).

**Table 7:** Correlations between RCSQ and hospitalization duration in days.

|  | **Hospitalization Days** | **p-value** |
| --- | --- | --- |
| 1. Sleep Depth | -0.293 | 0.001 |
| 2. Time to Fall Asleep | -0.434 | <0.001 |
| 3. Number of Awakenings | -0.379 | <0.001 |
| 4. Returning to Sleep | -0.296 | <0.001 |
| 5. Sleep Quality | -0.249 | 0.004 |
| 6. Noise Level | -0.370 | <0.001 |
| Total RCSQ (5 parameters) | -0.379 | <0.001 |

The duration of patients’ hospitalization under mechanical ventilation appeared to be negatively correlated with the RCSQ scales and the total RCSQ score (p<0.05). Slight and very slight statistically significant negative correlations were found, suggesting that as the duration of patients’ hospitalization under mechanical ventilation increases, their scores on the RCSQ scales and the total RCSQ score decrease (**Table 8**).

**Table 8:** Correlations between RCSQ and duration of mechanical ventilation in days.

|  | **Days on Mechanical Ventilation** | **p-value** |
| --- | --- | --- |
| 1. Sleep Depth | -0.298 | <0.001 |
| 2. Time to Fall Asleep | -0.437 | <0.001 |
| 3. Number of Awakenings | -0.373 | <0.001 |
| 4. Returning to Sleep | -0.297 | <0.001 |
| 5. Sleep Quality | -0.257 | 0.003 |
| 6. Noise Level | -0.376 | <0.001 |
| Total RCSQ (5 parameters) | -0.382 | <0.001 |

None of the RCSQ scales or the total RCSQ score were statistically significantly correlated with the duration of patients’ hospitalization under spontaneous breathing (p>0.05) (Table 9).

**Table 9:** Correlations between RCSQ and duration of spontaneous breathing in days.

|  | **Days on Spontaneous Breathing** | **p-value** |
| --- | --- | --- |
| 1. Sleep Depth | 0.002 | 0.983 |
| 2. Time to Fall Asleep | -0.103 | 0.238 |
| 3. Number of Awakenings | -0.147 | 0.091 |
| 4. Returning to Sleep | -0.064 | 0.463 |
| 5. Sleep Quality | 0.022 | 0.801 |
| 6. Noise Level | -0.010 | 0.911 |
| Total RCSQ (5 parameters) | -0.069 | 0.427 |

The scores on the RCSQ scales for Sleep Depth and Sleep Quality did not differ based on tracheotomy (p>0.05). In contrast, the scores on the RCSQ scales for Time to Fall Asleep, Number of Awakenings, Returning to Sleep, Noise Level, and the total RCSQ score were found to differ based on tracheotomy (p=0.007, p=0.008, p=0.015, p=0.022, and p=0.008, respectively).

Patients who underwent tracheotomy had lower scores on the Time to Fall Asleep, Number of Awakenings, Returning to Sleep, Noise Level scales, and the total RCSQ score compared to patients who did not undergo tracheotomy (**Table 10**).

**Table 10:** Comparisons between RCSQ and tracheotomy.

|  | **Tracheotomy** |  | **p-value** |
| --- | --- | --- | --- |
|  | No (n=59) | Yes (n=76) |  |
| 1. Sleep Depth | 57.97 ± 16.48 | 54.47 ± 13.99 | 0.080 |
| 2. Time to Fall Asleep | 53.22 ± 17.36 | 44.61 ± 18.86 | 0.007 |
| 3. Number of Awakenings | 50.34 ± 15.53 | 43.03 ± 16.00 | 0.008 |
| 4. Returning to Sleep | 55.25 ± 16.12 | 47.63 ± 18.68 | 0.015 |
| 5. Sleep Quality | 51.02 ± 17.78 | 44.21 ± 22.35 | 0.086 |
| 6. Noise Level | 55.76 ± 16.00 | 47.37 ± 20.87 | 0.022 |
| Total RCSQ (5 parameters) | 53.56 ± 14.33 | 46.79 ± 15.09 | 0.008 |

Finally, there was a statistically significant relationship between the RCSQ categories and the group to which the patients belonged (p<0.001). In the control group, most individuals (n=41) reported poor sleep, and none reported very good sleep. In contrast, in the intervention group, most individuals (n=54) reported good sleep, and 3 individuals reported very good sleep (Table 11 &12).

**Table 11:** Comparisons between RCSQ and patient group.

|  | Group |  | p-value |
| --- | --- | --- | --- |
|  | Control Group (n=62) | Intervention Group (n=73) |  |
| 1. Sleep Depth | 50.65 ± 15.35 | 60.55 ± 13.53 | <0.001 |
| 2. Time to Fall Asleep | 39.19 ± 18.40 | 56.16 ± 15.06 | <0.001 |
| 3. Number of Awakenings | 39.19 ± 15.92 | 52.19 ± 13.87 | <0.001 |
| 4. Returning to Sleep | 42.42 ± 17.43 | 58.22 ± 15.03 | <0.001 |
| 5. Sleep Quality | 35.65 ± 19.64 | 56.99 ± 16.05 | <0.001 |
| 6. Noise Level | 40.81 ± 18.13 | 59.73 ± 15.72 | <0.001 |
| Total RCSQ (5 parameters) | 41.42 ± 13.61 | 56.82 ± 12.51 | <0.001 |

**Table 12:**
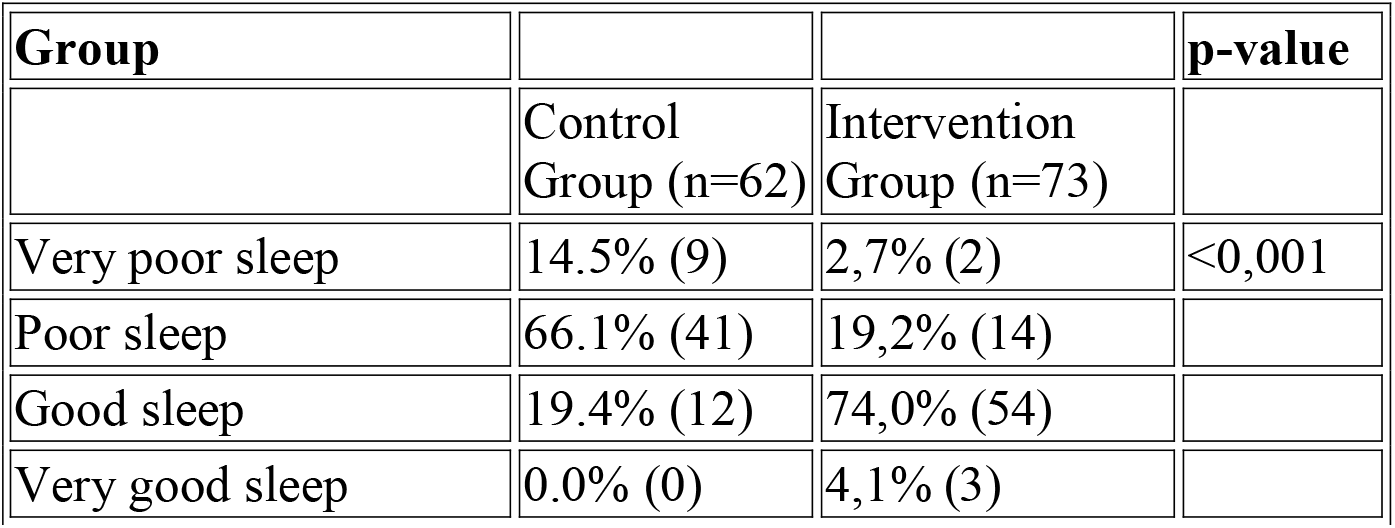
Correlations between RCSQ categories and patient group.

| Group |  |  | p-value |
| --- | --- | --- | --- |
|  | Control Group (n=62) | Intervention Group (n=73) |  |
| Very poor sleep | 14.5% (9) | 2,7% (2) | <0,001 |
| Poor sleep | 66.1% (41) | 19,2% (14) |  |
| Good sleep | 19.4% (12) | 74,0% (54) |  |
| Very good sleep | 0.0% (0) | 4,1% (3) |  |

## Discussion

Several studies have investigated sleep quality in ICU patients. A 2023 study by Ahn, Yoon Hae, et al., found that sleep disturbances are common in ICUs and identified several factors affecting sleep quality, such as noise, light, patient care interactions, physical discomfort, and illness. Physical discomfort, awakenings for interventions, and illness were significant barriers to sleep, with no correlation found between home sleep quality and ICU sleep quality. The Korean version of the Richards-Campbell Sleep Questionnaire (K-RCSQ) was used for subjective sleep quality assessment. The study identified modifiable factors that could improve sleep quality in ICU settings, emphasizing the need for further research in this area.^17^

The 2019 study by Lewandowska, Katarzyna, et al., aimed to evaluate sleep quality and the factors disrupting sleep in ICU patients in Northern Poland. Interviews with 83 ICU patients using the Short Portable Mental Status Questionnaire and the Richards-Campbell Sleep Questionnaire revealed that vital signs checks and light were the most disruptive sleep factors. Higher pain levels on the first ICU day were associated with greater sleep disruption. The study emphasizes the need for medical staff to be aware of and reduce sleep-disrupting factors.^18^

Naik, Ramavath Devendra, et al., aimed to assess the prevalence of poor sleep and identify factors affecting sleep quality in ICU patients. Actigraphy and the Richards-Campbell Sleep Questionnaire (RCSQ) were used in a cross-sectional study in a medical ICU. The findings revealed a high prevalence of poor sleep, particularly among mechanically ventilated patients, highlighting the need for non-pharmacological interventions to improve sleep quality in ICUs.^19^

A review published in Dove Press emphasized the importance of managing sleep disorders in ICUs. It noted that sleep disorders are prevalent in ICU patients due to various factors and may negatively impact patient outcomes. The review stressed the need for better sleep assessment tools and strategies to improve sleep quality in ICUs.^8^

These studies collectively highlight the importance of addressing sleep quality in ICU patients and the need for further research to develop effective strategies for improving sleep in this population.

Aydin Sayilan et al. (2021) aimed to examine the relationship between noise levels and pain, anxiety, and sleep quality in ICU patients. The study was conducted with 111 patients using sound level meters, pain and anxiety scales, and the Richards-Campbell Sleep Questionnaire. Noise levels in the ICU significantly exceeded WHO recommendations, affecting anxiety and sleep quality but not pain levels. The study highlights the need for noise reduction strategies in ICUs to improve patient outcomes.^20^

Noise is defined as unwanted sound and disturbance. Demir et al. (2017) aimed to study the impact of noise on sleep in intensive care patients and its effect on vital signs. The sample for this descriptive study was 83 patients hospitalized in the Neurosurgical ICU of Cukurova University Medical School, who met the study criteria, were over 18 years old, and agreed to participate. The study concluded that the average noise level was 52.04 ± 5.75 dB. Of the patients, 75% reported sleep problems due to noise, primarily from monitor alarms, with the most noticeable complaints being frequent awakenings. A slight positive correlation was found between noise level and systolic blood pressure, and a slight correlation with pulse, diastolic blood pressure, and respiratory noise level.^21^

Frisk, Ulla, and Gun Nordström aimed to describe ICU patients’ perceptions of their sleep and compare them with nurses’ perceptions. The Richards-Campbell Sleep Questionnaire (RCSQ) was used to assess sleep quality in 31 ICU patients. Patients who received hypnotics or sedatives reported significantly worse sleep. Noise and ICU procedures were not significant factors disrupting sleep. The RCSQ can be used by nurses to assess sleep in patients who cannot self-report, aiding in the evaluation of interventions to promote better sleep in ICU settings.^22^

The study by Simons, Koen S., et al. aimed to determine the effect of noise on sleep quality in ICU patients. This was a multicenter observational study in six Dutch ICUs with noise recording equipment installed in patient rooms. Environmental noise negatively affects sleep quality, while recovery periods and female gender are positively associated with better sleep.^23^

Menear, Ashika, et al. aimed to evaluate sleep quality in ICU patients using the Richards-Campbell Sleep Questionnaire (RCSQ) and the impact of sleep-promoting interventions. This was an observational study in a 58-bed adult ICU comparing current sleep-promoting intervention data with previous data. Despite the extensive use of sleep-promoting interventions, no significant improvement in sleep quality was observed following the implementation of a sleep guideline. The RCSQ is user-friendly for repeated assessment of sleep quality in ICU patients, but the effectiveness of sleep interventions needs further research.^24^

Another study aimed to describe ICU patients’ perceptions of their sleep quality and to develop a care plan to improve sleep. A prospective descriptive study was conducted with 125 patients using the Richards-Campbell Sleep Questionnaire and an ad-hoc questionnaire. Patients exhibited moderate sleep depth with light awakenings and difficulty falling back asleep.

Significant factors affecting sleep included pain, anxiety, staff voices, alarm sounds, and intravenous lines. Optimizing pain management, addressing concerns, and minimizing noise and voices could potentially improve sleep quality in ICU patients.^25^

In 2022, Pamuk, Kübra, and Nuray Turan aimed to assess the effect of light on sleep quality and physiological parameters in ICU patients. An experimental, randomized-controlled trial was conducted with 148 ICU patients, comparing a circadian lighting system with standard ICU lighting. The experimental group exposed to the circadian lighting system experienced significantly better sleep quality, with longer durations of deep and light sleep. The study concluded that ICU lighting affects sleep quality and physiological parameters, suggesting environmental adjustments to align with human circadian rhythms for improved outcomes.^26^

## Data Availability

All data produced in the present study are available upon reasonable request to the authors.

